# Two sample Mendelian Randomisation using an outcome from a multilevel model of disease progression

**DOI:** 10.1101/2023.04.27.23289203

**Authors:** Michael Lawton, Yoav Ben-Shlomo, Apostolos Gkatzionis, Michele T. Hu, Donald Grosset, Kate Tilling

## Abstract

Identifying factors that are causes of disease progression, especially in neurodegenerative diseases, is of considerable interest. Disease progression can be described as a trajectory of outcome over time - for example, a linear trajectory having both an intercept (severity at time zero) and a slope (rate of change). A technique for identifying causal relationships between one exposure and one outcome in observational data whilst avoiding bias due to confounding is two sample Mendelian Randomisation (2SMR). We consider a multivariate approach to 2SMR using a multilevel model for disease progression to estimate the causal effect an exposure has on the intercept and slope. We carry out a simulation study comparing a naïve univariate 2SMR approach to a multivariate 2SMR approach with one exposure that effects both the intercept and slope of an outcome that changes linearly with time since diagnosis. The simulation study results for both approaches were similar and approximately unbiased (bias for intercept ranges from -1.6% to 1.5% and the slope -0.7% to 4.1%) with appropriate coverage of the 95% confidence intervals (for intercept 94.1%-96.2% and the slope 94.7%-96.0%). The multivariate approach gives a better joint coverage of both the intercept and slope effects (93.3%-95.8% for multivariate approach compared to 89.0%-92.5% for the naïve approach). We also apply our method to two Parkinson’s cohorts to examine the effect body mass index has on disease progression. There was no strong evidence that BMI affects disease progression, however the confidence intervals for both intercept and slope were wide.

## 1. INTRODUCTION

Determining causality in observational cohort studies can be difficult due to problems with both measured and unmeasured confounding. Two sample Mendelian Randomisation (2SMR) is a technique used to determine causal relationships in observational studies that leverages genetic data as instrumental variables. Mendel’s laws state that genes are randomly assigned at conception hence they are ideal candidates for instrumental variables. Under the three instrumental variable assumptions 2SMR allows us to determine the causal relationship between an exposure and an outcome that is unaffected by confounding and reverse causation. This technique has gained popularity in recent years with the number of publications per year growing rapidly [1] and has mostly been used to determine relationships where the outcome is developing a disease (i.e. a binary outcome using logistic regression) [2, 3] or a health marker such as blood pressure (i.e. a continuous outcome using linear regression) [4-6]. Some research has also been carried out where the outcome is time to event [7], but to our knowledge not where the outcome is the trajectory of disease progression over time. We are interested in causal inference where the outcome is a repeatedly measured trait in individuals with a particular condition.

Neurodegenerative diseases like Parkinson’s disease (PD) and multiple sclerosis (MS) lead to disability that typically worsens over time. Identifying factors that are related to disease progression could lead to developing new treatments and better counselling of patients at diagnosis. Disease progression in observational cohorts has been studied before in both PD [8] and MS [9] using multilevel models (also called growth models, repeated measures models and random slope and intercept models). In these circumstances we are interested in the trajectory of some continuous trait over time which in the case of PD and MS is usually related to the severity of motor disability. When working with a multilevel model we are often interested in both the intercept (disability at a time of zero) and the slope (rate of change in disability over time) and it is important to study the effect of the exposure on both the intercept and slope.

This article presents a simulation study for a multivariate method to carry out 2SMR where we are interested in the causal effect an exposure has on both the intercept and slope in a model of disease progression. This method uses multivariate meta-analysis which is often used in meta-analysis of diagnostic studies when researchers are interested in both the sensitivity and specificity of a test [10, 11]. Our aim is to examine bias and coverage of both separate and joint confidence intervals using this approach. We also apply this method to two cohorts of individuals with PD where we are interested in the causal effect of body mass index (BMI) on severity at diagnosis (intercept) as well as disease progression (slope) using motor symptom severity measured by the Movement Disorder Society Unified Parkinson’s Disease Rating Scale (MDS-UPDRS) [12].

## 2. METHODS

### 2.1 Two sample Mendelian Randomisation

2SMR is a technique to estimate the causal effect (α) of an exposure on some outcome (y) using genetic data, usually single nucleotide polymorphisms (SNPs), as instrumental variables. The effect of each SNP on the exposure (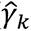 for the kth SNP) can be obtained from a genome-wide association study (GWAS). In the case of 2SMR the SNPs used should be independent. The effect of the SNPs on the outcome would come from a completely separate sample (hence the name two sample MR), for example they might come from some regression model with the following format.

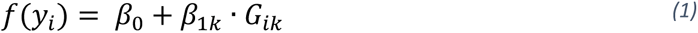

Where,

*G_ik_* = The number of effect alleles (0, 1 or 2) for the *i*th individual and *k*th SNP

*y_i_* = Outcome for the *i*th individual

Using the data from these two samples the causal effect can be estimated from a weighted regression of the estimated effects of SNPs on the outcome (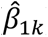) against the estimated effects of SNPs on the exposure (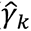). This is weighted by the inverse of the variance of the effects of SNPs on the outcome (*var* 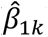)).

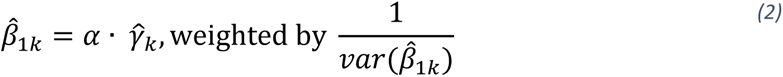

This can also be thought of as a meta-regression or a meta-analysis of Wald ratios (in this example 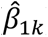/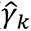) which are all identical mathematically. There are other approaches to 2SMR which relax the assumptions in some way the most common being the MR-Egger [13], Median [14], and Mode [15] approaches. None of these approaches require that the gene-exposure association is linear, or that the model for the exposure is correctly specified [16].

### 2.2 Disease progression models

We are focussing here on multilevel models (also called growth models, random slope and intercept models and hierarchical models). They are often used to model outcomes across time where repeated measurements are available and can easily accommodate unbalanced data (number of observations per individual differs and the time between observations is not constant). They account for the non-independent nature of repeated measurements within an individual by incorporating random effects into a standard regression model. A simple model with no covariates other than time where the relationship with time was linear would have the format

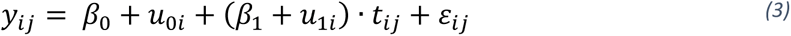

*i = 1, …, n* (number of individuals)

*j = 1, …, n_j_* (number of observations per person)

*t_ij_ =* time at the *j*th time point for the *i*th individual

*y_ij_ =* outcome at the *j*th time point for the *i*th individual

ε_*ij*_∼*N*[0,σ^2^] - this is the residual variation, also sometimes called level 1 variation in context of multilevel models assumed to be normally distributed

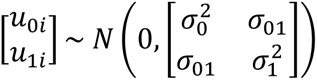 – these are the patient level random effects, assumed to be bivariate normally distributed.

For simplicity in this paper we are assuming models with a continuous outcome, only linear time, no complex level 1 variation, and no informative drop-out however these models could easily be adapted as explained later in the discussion. Within this paper we are assuming a particular directed acyclic graph (DAG), see figure 1. That is the intercept does not cause the slope but the exposure causes some latent progression trait that causes both the intercept and slope.

**Figure 1.**
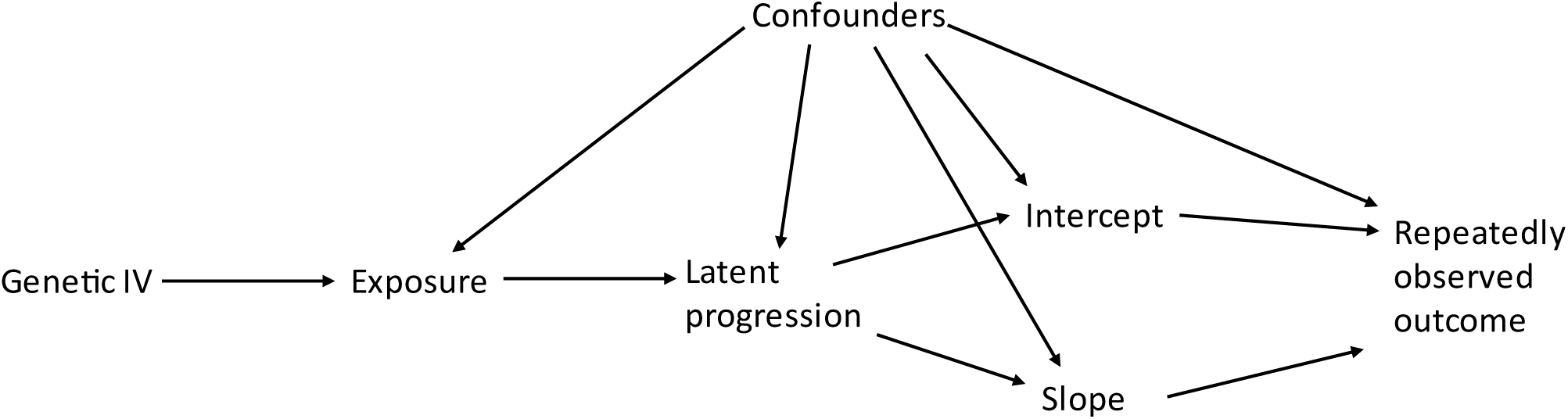
Assumed Directed Acyclic Graph for the effect of exposure on disease progression.

In the context of causal effect modelling and the DAG we would be interested in the effect that some exposure had on both the intercept and the slope. If there was an effect on the intercept but not the slope then we could say that the exposure was related to disease severity at baseline and if there was an effect on the slope that the exposure was related to disease progression (or rate of change). If there was a single confounder of the effect the exposure has on the intercept and slope we would have the following longitudinal model

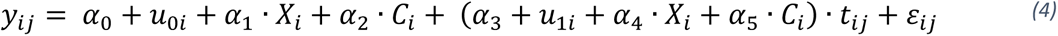

*X_i_ =* the exposure for the *i*th individual

*C_i_ =* the confounder for the *i*th individual

Here α_1_ is the causal effect of the exposure on the intercept and α_4_ the causal effect of the exposure on the slope.

In the context of 2SMR we would then be fitting a multilevel model for each SNP such that

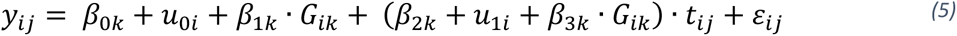

and from the GWAS study we would know (in the case of a continuous exposure with linear regression) that for *k* SNPs

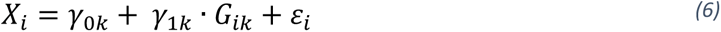

If we insert equation 6 into equation 4 and compare the with equation 5 we can see that

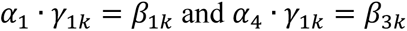

A naïve approach to 2SMR would be to carry out a meta-regression of the 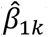 ‘s and 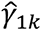 ’s to estimate α_1_ the causal effect of the exposure on the intercept. In a standard approach to 2SMR this would be a fixed-effects meta-regression weighted by the inverse of the variance of the 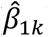 ‘s. Under the assumption that these SNPs are independent then meta-regression would be a valid method to estimate α_1_.

Then we could carry out a separate meta-regression of the 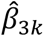 ‘s and 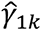 ’s to estimate α_4_ the causal effect of the exposure on the slope. Again this would be a fixed-effects meta-regression weighted by the inverse of the variance of the 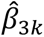 ‘s.

This would be complicated by each SNP having an effect on both the intercept and the slope and those effects could be correlated. Hence there is a covariance between the 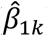 ’s and the 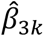 ’s whilst α_1_ and α_4_ could also be correlated. Using multivariate meta-regression [17] we could estimate both α_1_and α_4_ jointly incorporating the covariance between the 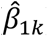 ’s and the 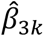 ’s. Multivariate meta-analysis and meta-regression is a likelihood based method that applies weights to the likelihood using the covariance matrix of the 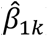 ’s and the 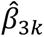 ’s. To be consistent with standard 2SMR we have used a fixed effects estimation. Doing the estimation jointly allows us to estimate not only the effect an exposure has on the intercept and on the slope (along with the standard errors) but also the covariance between these two effects.

### 2.3 Application of approach to Parkinson’s cohort

We will motivate this 2SMR approach using two parallel cohorts of individuals with PD, the Oxford Discovery cohort and the Tracking Parkinson’s cohort [18, 19]. At recruitment individuals had to be within 3.5 years of diagnosis and are followed up every 18 months in both studies. The Oxford Discovery cohort was recruited from 11 hospitals in the Thames Valley Region between 2010 and 2016. The Tracking cohort was recruited from 72 sites across the UK between 2012 and 2014. We have previously carried out a genome-wide association study of motor and cognitive progression in these two cohorts [20]. Our analysis samples will be restricted to those with a probability of diagnosis of PD ≥ 90% as rated by a neurologist at the latest available visit. This is an attempt to exclude individuals who were incorrectly diagnosed with PD as it has been shown previously that some individuals diagnosed with PD will turn out to have another disorder [21, 22]. More details about the individuals are included in the results. Informed consent was obtained from all individual participants included in both studies.

The outcome (MDS-UPDRS part III) we are using consists of 33 questions rated on a scale of 0-4 giving a total score that ranges from 0-132 [12]. This is the most common instrument for motor symptom severity within the field of PD and is often used as the primary outcome in RCTs. For people with PD it does not exhibit a floor or ceiling effect and although technically measured at an ordinal scale, its range is large enough that it can be considered approximately continuous. We are using time since diagnosis as the time axis in our multilevel models.

### 2.4 Simulation study

For our simulation study we have used the ADEMP (aim, data, estimands, methods, performance) guidelines to inform the design and reporting [23].

#### Aim

The aim of this study was to investigate different methods for two sample Mendelian Randomisation where the outcome is a multilevel model of disease progression.

#### Data generating mechanism

We simulated data for 10,000 individuals. Our simulation was partially informed by the subsequent real-data application studying the effects of body mass index (BMI) on Parkinson’s disease. We generated genetic data based on the Locke BMI GWAS paper [24]. This paper reported on 97 GWAS hits. The number of effect alleles for each individual was simulated from a binomial distribution with n = 2 and p = effect allele frequency from the BMI GWAS paper. Using the beta-coefficients reported from the BMI GWAS paper we were able to create an exposure measurement for each individual by multiplying the simulated SNPs by their beta-coefficients (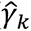) and then adding on an additional residual variance term. We also simulated a continuous confounder (for the exposure and outcome) that would describe 50% of this residual variance in the exposure. This variance term and confounder were simulated for both an R^2^ of 2% and 10% to assess whether the variance explained by our genetic instruments affected the methods performance. To construct this variance term we calculated the expected variance of the 97 SNPs by the sum of 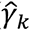^2^*2*p*(1-p). Note that in the BMI GWAS paper the SNPs actually explained 2.7% of the variance of BMI.

We then simulated balanced longitudinal data with 7 visits per person observed at times of 0, 1, … to 6. The data was simulated under four different scenarios described below.

After simulating the SNPs, the exposure and the confounder the longitudinal data was simulated under the following model with the same definitions as in equation 1 above.

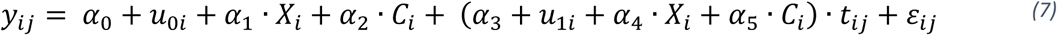

##### Fixed effects

In scenarios 1, 3 and 4 we used the following parameter estimates.

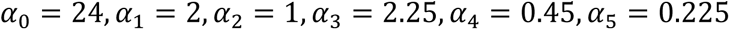

In scenario 2 we altered the effect the exposure and confounder had on the slope to change the relative effect of intercept vs. slope, such that

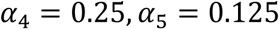

The effect the exposure and confounder had on the intercept (α_1_, α_2_) and the slope (α_4_, α_5_) was based on the expected s.d. of the exposure and confounder. This was to ensure similarity of effect sizes when R^2^ was 2% and 10% since changing the variance term when simulating the exposure would also alter the variance of the exposure. Note the direct effect the confounder has on the intercept and slope was 50% of the total effect the exposure had, however there is also a substantial indirect effect as the confounder described 50% of the residual variance in the exposure.

#### Random effects and residuals

For scenarios 1 and 2 the patient level random effects were simulated from

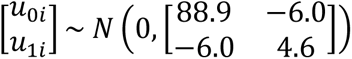

In the third and fourth scenarios we altered the covariance of the random effects thus allowing us to alter the estimated covariance between the SNP-intercept and SNP-slope effects. In scenario 3 the covariance was adjusted to be positive (6.0) and in scenario 4 the covariance was adjusted to be twice as large as scenario 3 (12.0). The level 1 residuals were simulated from

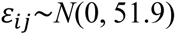

The random effects distribution (for scenarios 1 and 2) and the level 1 residual term were taken from an analysis on the PD Discovery cohort MDS-UPDRS III data (see section 2.4 below).

Finally we carried out a sensitivity analysis with a smaller sample size and unbalanced data with fewer observations. This used the same approach as scenario 1 with an R^2^ of 10% but only 1,000 individuals (instead of 10,000) and unbalanced data. To create the unbalanced data each individual had 1 observation with 20% probability, 2 observations with 30% probability, 3 observations with 30% probability or 4 with 20% probability. The baseline time was simulated from a uniform distribution between 0 and 3.5 and the time between each observation was simulated from a uniform distribution between 1.4 and 1.6.

For each scenario, 1,000 simulations were run.

#### Estimands/targets of analysis

The estimand of interest is the effect the exposure (BMI) has on the intercept (α_1_) and also the slope (α_4_) in the model of disease progression (equation 4).

#### Methods

For each SNP we fit a multilevel model of the following format

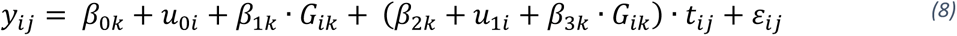

All other variables are as described above in section 2.2.

After fitting one multilevel for each SNP we considered two different approaches. The first is a naïve approach doing two separate 2SMR’s as described above in section 2.2 one for the intercept (using 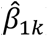 ′s) the other for the slope (using 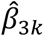 ′s).

The second approach is a multivariate approach using multivariate meta-analysis techniques [17] which allows us to also incorporate the estimated covariance between 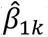 and 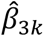 as described above in section 2.2

#### Performance metrics

We are primarily interested in the bias and coverage of the 95% confidence intervals of the exposure-intercept and exposure-slope effects and also report the empirical SD and the mean of the model based standard errors. Finally we report the joint coverage of the confidence intervals for both the exposure-intercept and exposure-slope effect estimates. The multivariate approach can be used to construct a joint confidence region for the intercept and the slope; such a region will have an elliptic shape, as opposed to a box-shaped region that is obtained by combining the two separate confidence intervals produced by the naïve approach.

### 2.5 Computing

This work was carried out using the computational facilities of the Advanced Computing Research Centre, University of Bristol – http://www.bristol.ac.uk/acrc/. All the simulations and analyses were carried out within STATA 17 and we used the mvmeta package for the multivariate meta-analysis [17].

## 3 RESULTS

### 3.1 Motivating example in Parkinson’s Disease

The Discovery cohort analysis is based on 826 individuals with 2,851 observations (average 3.5 ranging from 1 to 7) over an average of 4.2 years follow-up. The average age at diagnosis was 66.0 years (SD 9.6 years) with 538 (65.1%) males. Average disease duration at baseline was 1.2 years (SD 0.9). In a multilevel model where the MDS-UPDRS III was the outcome the fixed effect for the intercept was 23.5 (95% CI: 22.6 to 24.3) and the fixed effect for the linear slope was 2.36 (95% CI: 2.14 to 2.60), i.e. the average outcome at diagnosis was 23.5 and the outcome increased at 2.36 points per year after diagnosis.

The Tracking cohort is based on 1,517 individuals with 5,024 observations (average 3.3 ranging from 1 to 6) over an average of 3.8 years of follow-up. The average age at diagnosis was 65.9 years (SD 9.2 years) with 987 (65.1%) males which is almost identical to the Discovery cohort. Average disease duration at baseline was 1.3 years (SD 0.9). In a multilevel model where the MDS-UPDRS III was the outcome the fixed effect for the intercept was 20.4 (95% CI: 19.6 to 21.1) and the fixed effect for the linear slope was 2.24 (95% CI: 2.06 to 2.43).

In table 1 we report the estimates for the effect the exposure has on the intercept and the slope within the two Parkinson’s cohorts and a meta-analysis of the two cohorts. The meta-analysis of the two cohorts for the naïve approach comes from two fixed effects meta-analyses (one for the slope and another for the intercept. Whilst the meta-analysis of the two cohorts for the multivariate approach comes from a multivariate meta-analysis. The results from the naïve and multivariate approaches are very similar. There is a large degree of uncertainty in these estimates (especially when compare to the average intercept and slope reported above) and despite the fact the estimates of the effect of exposure (BMI) on slope are in opposite directions in the two cohorts the 95% confidence intervals overlap with each other. Given the uncertainty, which is probably due to the limited sample size (for genetic studies) along with the relatively small genetic effects, it is difficult to rule out either a protective or detrimental effect of BMI on disease progression. Figure 2 (meta-analysis) and supplementary figure 1 (cohort specific effects) clearly shows that when plotting the effect that the exposure has on the outcome across time that using the naïve approach (and thus ignoring the covariance) there is a considerable difference to the confidence intervals.

**Table 1.** Example 2SMR analysis on the Discovery and Tracking cohorts with BMI as the exposure and MDS-UPDRS III^a^ as the outcome. Data is estimate (95% CI); p-value

|  | Discovery cohort |  | Tracking cohort |  | Meta-analysis of two cohorts<sup>b</sup> |  |
| --- | --- | --- | --- | --- | --- | --- |
| Analysis | Intercept | Slope | Intercept | Slope | Intercept | Slope |
| naïve approach | -2.43<br>(-7.66, 2.79);<br>p=0.36 | 0.385<br>(-1.014, 1.784);<br>p=0.59 | -0.086<br>(-4.41, 4.24);<br>p=0.97 | -0.169<br>(-1.28, 0.94);<br>p=0.77 | -1.04<br>(-4.37, 2.29);<br>p=0.54 | 0.045<br>(-0.83, 0.92);<br>p=0.92 |
| multivariate approach | -2.47<br>(-7.69, 2.75);<br>p=0.35 | 0.389<br>(-1.011, 1.788);<br>p=0.59 | -0.065<br>(-4.39, 4.26);<br>p=0.98 | -0.165<br>(-1.28, 0.95);<br>p=0.77 | -1.04<br>(-4.37, 2.29);<br>p=0.54 | 0.054<br>(-0.82, 0.92);<br>p=0.90 |
|  | Estimated (multivariate approach)<br>correlation = -0.52 |  | Estimated (multivariate approach)<br>correlation = -0.54 |  | Estimated (multivariate approach) correlation<br>= -0.53 |  |

<sup>a</sup>Note that higher scores on the MDS-UPDRS III are worse symptoms. So, for example, a positive association with the intercept would reflect higher BMI being associated with worse symptoms at baseline.
<sup>b</sup>Within the naïve approach to 2SMR the intercept and slope for the two cohorts are meta-analysed with a standard univariate approach. For the multivariate approach to the 2SMR the intercept and slope for the two cohorts are meta-analysed with a multivariate approach accounting for estimated the covariance.

**Figure 2.**
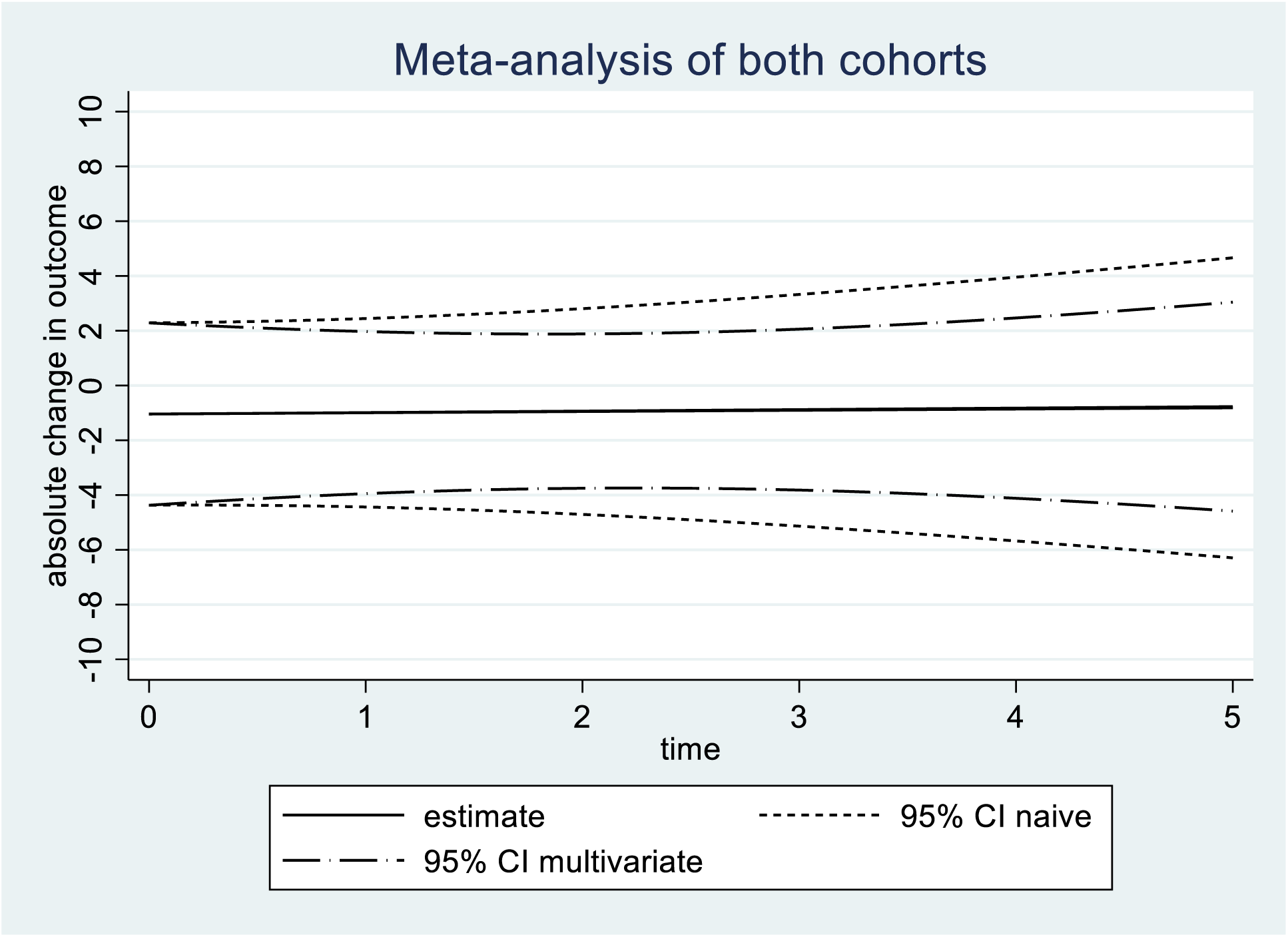
Absolute change in the MDS-UPDRS III for a one point increase in the exposure. Estimated model from the meta-analysis of the two cohorts including the 95% confidence intervals for both the naïve and multivariate approach

### 3.2 Simulation study

The results from our simulation with R^2^ = 10% are displayed in table 2. The results based on the naïve and multivariate approach were almost identical in this simulation and you generally had to look to the third significant figure to see any differences. In the four different scenarios the bias for the intercept ranged from -0.2% to 0.3% and for the slope from -0.3% to 0.7%. The coverage of the 95% confidence intervals were also at nominal levels: the coverage for the intercept ranged from 94.2% to 96.1% and for the slope from 94.7% to 95.8%.

**Table 2.** Results from the simulation based on R^2^ of 10%

|  |  | Intercept – naïve approach |  |  |  | Intercept – multivariate approach |  |  |  |
| --- | --- | --- | --- | --- | --- | --- | --- | --- | --- |
| Scenario (truth) | Correlation<sup>a</sup> mean (sd) | Estimate mean (sd) | Mean model se | Coverage 95% CI | Mean relative Bias<sup>b</sup> | Estimate mean (sd) | Mean model based se | Coverage 95% CI | Mean Relative Bias<sup>b</sup> |
| 1 (2) | -0.35 (0.009) | 2.00 (0.338) | 0.341 | 94.8% | -0.2% | 2.00 (0.338) | 0.341 | 94.8% | -0.2% |
| 2 (2) | -0.38 (0.008) | 2.01 (0.349) | 0.341 | 94.7% | 0.3% | 2.01 (0.349) | 0.341 | 94.7% | 0.3% |
| 3 (2) | 0.076 (0.010) | 2.00 (0.347) | 0.341 | 94.2% | 0.1% | 2.00 (0.347) | 0.341 | 94.2% | 0.1% |
| 4 (2) | 0.29 (0.009) | 2.00 (0.328) | 0.341 | 96.1% | -0.2% | 2.00 (0.328) | 0.341 | 96.1% | -0.2% |
|  |  | Slope – naïve approach |  |  |  | Slope – multivariate approach |  |  |  |
| Scenario (truth) | Correlation (sd) | Estimate mean (sd) | Mean model based se | Coverage 95% CI | Mean relative Bias<sup>b</sup> | Estimate mean (sd) | Mean model based se | Coverage 95% CI | Mean relative Bias<sup>b</sup> |
| 1 (0.45) | -0.35 (0.009) | 0.449 (0.081) | 0.083 | 95.8% | -0.3% | 0.449 (0.081) | 0.083 | 95.8% | -0.3% |
| 2 (0.25) | -0.38 (0.008) | 0.251 (0.080) | 0.081 | 95.4% | 0.3% | 0.251 (0.080) | 0.081 | 95.4% | 0.3% |
| 3 (0.45) | 0.076 (0.010) | 0.453 (0.081) | 0.083 | 94.7% | 0.7% | 0.453 (0.081) | 0.083 | 94.7% | 0.7% |
| 4 (0.45) | 0.29 (0.009) | 0.449 (0.083) | 0.083 | 94.7% | -0.2% | 0.449 (0.083) | 0.083 | 94.7% | -0.2% |

<sup>a</sup>Estimated average correlation from the covariance of SNP-intercept and SNP-slope effects
<sup>b</sup>(observed estimate-true)/true

The results from our simulation with R^2^ = 2%, displayed in table 3, were similar. The bias for the intercept ranged from -1.6% to 1.5% and for the slope from -0.7% to 4.1%. The coverage of the 95% confidence interval for the intercept ranged from 94.1% to 96.2% and for the slope from 95.0% to 96.0%.

**Table 3.**
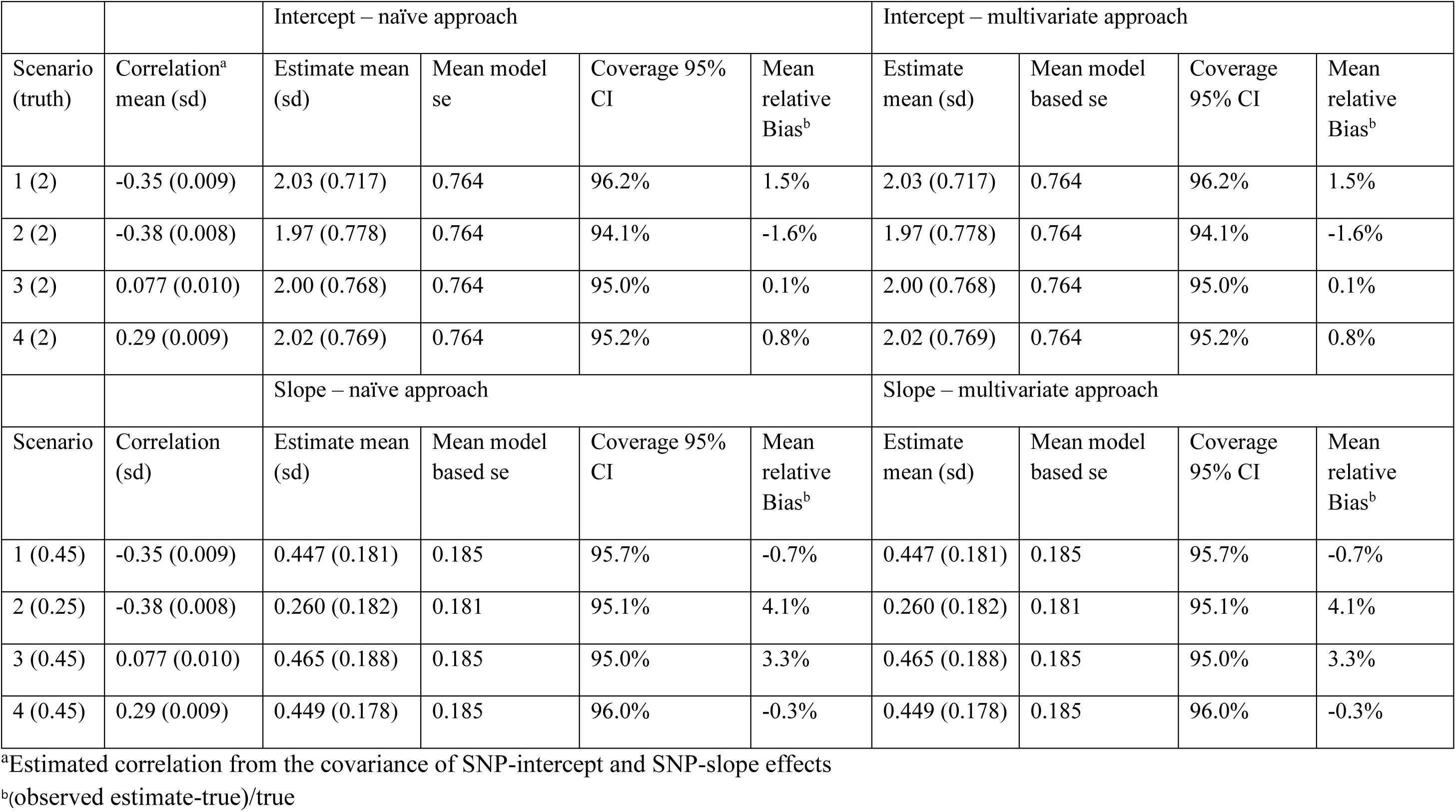
. Results from the simulation based on R^2^ of 2%

|  |  | Intercept – naïve approach |  |  |  | Intercept – multivariate approach |  |  |  |
| --- | --- | --- | --- | --- | --- | --- | --- | --- | --- |
| Scenario (truth) | Correlation<sup>a</sup> mean (sd) | Estimate mean (sd) | Mean model se | Coverage 95% CI | Mean relative Bias<sup>b</sup> | Estimate mean (sd) | Mean model based se | Coverage 95% CI | Mean relative Bias<sup>b</sup> |
| 1 (2) | -0.35 (0.009) | 2.03 (0.717) | 0.764 | 96.2% | 1.5% | 2.03 (0.717) | 0.764 | 96.2% | 1.5% |
| 2 (2) | -0.38 (0.008) | 1.97 (0.778) | 0.764 | 94.1% | -1.6% | 1.97 (0.778) | 0.764 | 94.1% | -1.6% |
| 3 (2) | 0.077 (0.010) | 2.00 (0.768) | 0.764 | 95.0% | 0.1% | 2.00 (0.768) | 0.764 | 95.0% | 0.1% |
| 4 (2) | 0.29 (0.009) | 2.02 (0.769) | 0.764 | 95.2% | 0.8% | 2.02 (0.769) | 0.764 | 95.2% | 0.8% |
|  |  | Slope – naïve approach |  |  |  | Slope – multivariate approach |  |  |  |
| Scenario | Correlation (sd) | Estimate mean (sd) | Mean model based se | Coverage 95% CI | Mean relative Bias<sup>b</sup> | Estimate mean (sd) | Mean model based se | Coverage 95% CI | Mean relative Bias<sup>b</sup> |
| 1 (0.45) | -0.35 (0.009) | 0.447 (0.181) | 0.185 | 95.7% | -0.7% | 0.447 (0.181) | 0.185 | 95.7% | -0.7% |
| 2 (0.25) | -0.38 (0.008) | 0.260 (0.182) | 0.181 | 95.1% | 4.1% | 0.260 (0.182) | 0.181 | 95.1% | 4.1% |
| 3 (0.45) | 0.077 (0.010) | 0.465 (0.188) | 0.185 | 95.0% | 3.3% | 0.465 (0.188) | 0.185 | 95.0% | 3.3% |
| 4 (0.45) | 0.29 (0.009) | 0.449 (0.178) | 0.185 | 96.0% | -0.3% | 0.449 (0.178) | 0.185 | 96.0% | -0.3% |

<sup>a</sup>Estimated correlation from the covariance of SNP-intercept and SNP-slope effects
<sup>b</sup>(observed estimate-true)/true

Table 4 shows the joint coverage for the two approaches. For the naïve approach the joint coverage ranged from 89.0% to 91.5% when the R^2^ was 10% and from 89.9% to 92.5% when the R^2^ was 2%. This is not surprising, as the joint coverage of two separate 95% confidence intervals is approximately 0.95^2 = 0.9025. Using the multivariate approach the joint coverage increased and ranged from 94.6% to 95.2% when the R^2^ was 10% and from 93.3% to 95.8% when the R^2^ was 2%.

**Table 4.** Joint coverage of the intercept and slope. For the naïve approach we used the two confidence intervals (essentially drawing a box) whilst for the multivariate approach we have drawn a confidence ellipse.

|  | R<sup>2</sup> of 10% |  | R<sup>2</sup> of 2% |  |
| --- | --- | --- | --- | --- |
| Scenario | Coverage of naïve approach 95% CI | Coverage of multivariate approach 95% CI | Coverage of naïve approach 95% CI | Coverage of multivariate approach 95% CI |
| 1 | 91.5% | 94.9% | 92.5% | 95.5% |
| 2 | 90.6% | 94.9% | 89.9% | 93.3% |
| 3 | 89.0% | 94.6% | 90.5% | 94.5% |
| 4 | 91.3% | 95.2% | 91.5% | 95.8% |

In our sensitivity analysis, see supplementary table 1, with a much reduced sample size and unbalanced data the naïve and multivariate approach still gave near identical results. Not surprisingly the uncertainty in the estimates is much larger than observed in tables 2 and 3. The bias was still small approximately -0.15% for the intercept and approximately -1.35% for the slope. The coverage of the 95% CI was 94.8% and 95.0% for the intercept (using the naïve and multivariate approach respectively) and 94.4% and 94.3% for the slope (using the naïve and multivariate approach respectively). The joint coverage for the sensitivity analysis was 92.5% for the naïve approach and 94.6% for the multivariate.

## 4. DISCUSSION

Our simulations show that 2SMR gives unbiased estimates with good coverage when the outcome is a multilevel model of linear disease progression. If an individual is interested in graphing the effect an exposure will have on an outcome over time then a multivariate meta-analysis method that allows the estimation of a covariance term could give substantially different confidence intervals. The multivariate meta-analysis method also allows for appropriate joint coverage of both the intercept and slope effects by drawing a confidence ellipse.

In practice progression trajectories might be non-linear in time [9] and in these cases we would recommend deriving the difference in trajectories per additional copy of the effect allele at some time-point. The chosen time-point will depend on both the length of follow-up in the data and what would be a meaningful time since onset/diagnosis in that particular disease. Our multivariate meta-analysis approach could also easily be adapted to have multiple time terms and could be used to create a graph similar to that displayed in figure 2. Non-linearity could also be a problem in our PD data however the number of observations we have per-person (on average) is limited so there is little data to test and account for non-linear trajectories.

The models we have used assume that there is measurement error in each observed measure of outcome, but these errors are unrelated and the variance is constant over time. The multilevel models can be modified to allow autocorrelation, or complex measurement error where the variance of the level 1 residuals changes over time [9]. If there is an underlying factor affecting measurement error in all measures for an individual – e.g. if individuals with impaired hearing tend to always score lower due to difficulties completing the questionnaire – then the measurement errors for all the measures will be correlated. However, this correlation will be subsumed into the effect of the “latent progression” factor, and thus also included in the model-based estimate of correlation between intercept and slope. This type of measurement error would not bias the MR in this example, because hearing loss would be essentially a confounder of different measures of outcome [25].

Other problems in observational longitudinal data occur when the follow-up data is missing not at random (informative drop-out), for example where an individual’s disability becomes so severe they can no longer attend clinic visits which could bias trajectories towards the null. This could be explored using pattern-mixture models, selection models or other similar approaches [26-29] that under some untestable assumptions would be less biased than standard multilevel models. We have used pattern-mixture models before to look at disease progression in PD which gave us similar results to a standard multi-level model providing some evidence that informative drop-out did not bias our results [8].

Allowing an exposure to have an effect on both the intercept and slope is possibly an over-simplistic way to model progression in PD. In reality there should be a true time zero where the individual has no impairment caused by the disease. However in PD there is a long prodromal (prediagnostic) period [30] and by the time of diagnosis there is already considerable motor impairment and large variability in motor impairment between individuals [31]. Modelling the effect of the exposure on the intercept using disease duration from diagnosis as the time-axis reflects some period of prodromal progression. The current progression data that is available for most cohorts of individuals with PD does not allow us to observe this prodromal period.

Another problem with carrying out 2SMR in a cohort of individuals who have disease is the index event bias phenomenon. Index event bias occurs when there are confounders of incidence and prognosis. By conditioning on individuals who have developed the disease we condition on a collider and can induce bias which is explained in more detail here [32]. In the case of 2SMR, two methods to address this bias are Dubridge et al’s index event bias correction [33] and Slope-Hunter [34] which were both originally developed for genome wide association studies and would also require additional data from a genome-wide association study of developing the disease in question. A recent review of methods for addressing index event bias was conducted by Mitchell et al. [35] and we refer you to these papers for more details.

A 2SMR package in STATA [36] by default forces the residual variance to be 1 when residual variance is less than one or freely estimates the residual variance when it is greater than one, thus allowing for overdispersion but not underdispersion. Not allowing for underdispersion is intuitive since we are weighting by the inverse of the variance of each SNP and it would be strange to allow for an estimator that is more precise than the variability in our SNP-outcome measures. Unfortunately our multivariate meta-analysis approach will not allow for overdispersion so is the same as forcing the residual variance to be 1. If there was overdispersion then our method could potentially result in an estimate that is overly precise.

Our approach is very similar in nature to a two-stage multivariate MR method for mixed outcomes (called MRMO) [37] which studies the effect of an exposure on multiple outcomes (which could be mixed such as binary and continuous). MRMO also describes a procedure to estimate a p-value for whether the exposure has an effect on any of the outcomes by using a Wald test. We could also estimate a Wald test using our estimates for the exposure-intercept and exposure-slope along with their covariance matrix. This would be a joint hypothesis test for whether the exposure-intercept is zero and the exposure-slope is zero and could be interpreted as whether the exposure has any effect on progression. However we would prefer to focus on confidence intervals and regions rather than p-values.

Using our multivariate meta-analysis method it would be possible to carry out the MR-Egger approach by also estimating an intercept term allowing us to correct for directional pleiotropy. In future work we will explore whether other methods such as the median and mode approaches could be adapted for this purpose. Given the large uncertainty from the analysis presented here, large consortiums with many longitudinal PD cohorts will be required to have sufficient power to detect whether exposures are related to progression.

This paper shows that is possible to use multivariate meta-analysis to carry out two sample Mendelian Randomisation when using an outcome that is repeatedly measured over time. The associations between an exposure and the intercept and linear slope of the repeatedly measure trait are unbiased with valid confidence intervals. We hope this inspires more people to use two sample Mendelian Randomisation to test hypotheses about exposures being related to disease progression in neurodegenerative diseases.

## Data Availability

Data from the Oxford Discovery cohort is available on request from https://www.dpag.ox.ac.uk/opdc/research/external-collaborations
Data from the Tracking Parkinsons cohort is available on request from https://www.trackingparkinsons.org.uk/about-1/data/

## ETHICS APPROVAL STATEMENT

The Oxford Discovery cohort was approved by NRES Committee, South Central Oxford A Research Ethics Committee, Reference number 16/SC/0108

The Tracking Parkinsons cohort was approved by West of Scotland Research Ethics Service (WoSRES) reference 11/AL/0163

## DATA SHARING STATEMENT

Data from the Oxford Discovery cohort is available on request from https://www.dpag.ox.ac.uk/opdc/research/external-collaborations

Data from the Tracking Parkinsons cohort is available on request from https://www.trackingparkinsons.org.uk/about-1/data/

## AUTHOR CONTRIBUTIONS STATEMENT

Michael Lawton - study concept and design, analysis and interpretation of the data, writing of the manuscript

Yoav Ben-Shlomo - study concept and design, revision of the manuscript

Apostolos Gkatzionis - study concept and design, analysis and interpretation of the data, revision of the manuscript

Michele T.M. Hu - study concept and design, acquisition of data, revision of the manuscript Donald Grosset - study concept and design, acquisition of data, revision of the manuscript Kate Tilling - study concept and design, analysis and interpretation of the data, revision of the manuscript

## ACKNOWLEDGEMENTS

The Oxford Discovery study was funded by the Monument Trust Discovery Award from Parkinson’s UK and supported by the National Institute for Health Research (NIHR) Oxford Biomedical Research Centre based at Oxford University Hospitals NHS Trust and University of Oxford, and the NIHR Clinical Research Network: Thames Valley and South Midlands

The Tracking Parkinson’s study was funded by Parkinson’s UK and supported by the National Institute for Health Research (NIHR) DeNDRoN network, the NIHR Newcastle Biomedical Research Unit based at Newcastle upon Tyne Hospitals NHS Foundation Trust and Newcastle University, and the NIHR funded Biomedical Research Centre in Cambridge (Grant number:146281) . The views expressed are those of the authors and not necessarily those of the NHS, the NIHR or the Department of Health.

---

**Supplementary Table 1.** Results from the simulation with a reduced sample size and unbalanced data (between 1-4 observations per person)

|  |  | Intercept – naïve approach |  |  |  | Intercept – multivariate approach |  |  |  |
| --- | --- | --- | --- | --- | --- | --- | --- | --- | --- |
| Scenario (truth) | Correlation<sup>a</sup> mean (sd) | Estimate mean (sd) | Mean model se | Coverage 95% CI | Mean relative Bias<sup>b</sup> | Estimate mean (sd) | Mean model based se | Coverage 95% CI | Mean relative Bias<sup>b</sup> |
| 1 (2) | -0.65 (0.019) | 2.00 (1.54) | 1.52 | 94.8% | -0.2% | 2.00 (1.54) | 1.52 | 95.0% | -0.1% |
|  |  | Slope – naïve approach |  |  |  | Slope – multivariate approach |  |  |  |
| Scenario | Correlation (sd) | Estimate mean (sd) | Mean model based se | Coverage 95% CI | Mean relative Bias<sup>b</sup> | Estimate mean (sd) | Mean model based se | Coverage 95% CI | Mean relative Bias<sup>b</sup> |
| 1 (0.45) | -0.65 (0.019) | 0.444 (0.454) | 0.446 | 94.4% | -1.4% | 0.444 (0.454) | 0.446 | 94.3% | -1.3% |
|  |  | Joint coverage - naïve approach |  |  |  | Joint coverage - multivariate approach |  |  |  |
|  |  | 92.5% |  |  |  | 94.6% |  |  |  |

<sup>a</sup>Estimated correlation from the covariance of SNP-intercept and SNP-slope effects
<sup>b</sup>(observed estimate-true)/true

**Supplementary Figure 1.**
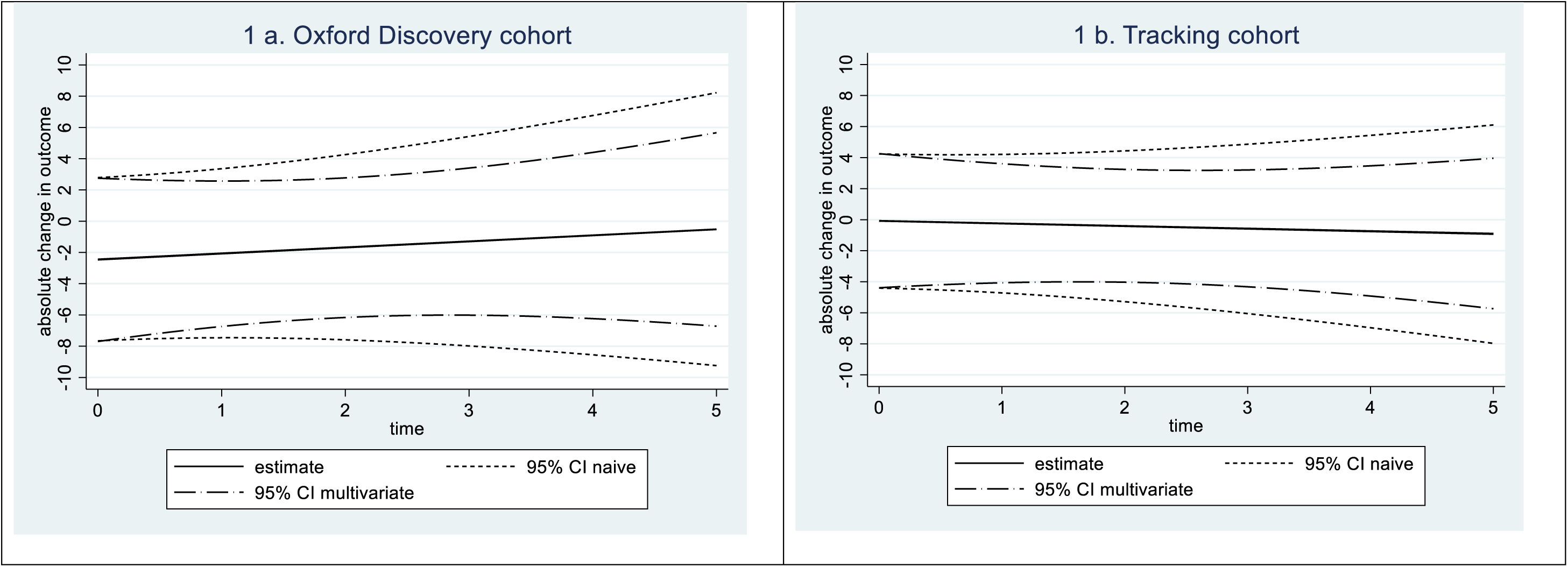
Absolute change in the MDS-UPDRS III for a one point increase in the exposure. 1a shows the estimated model from the Oxford Discovery cohort including the 95% confidence intervals for both the naïve and multivariate approach and 1b the same from the Tracking cohort.

